# Increasing SARS-CoV-2 RT-qPCR testing capacity by sample pooling

**DOI:** 10.1101/2020.10.24.20218685

**Authors:** Julia Alcoba-Florez, Helena Gil-Campesino, Diego García-Martínez de Artola, Oscar Díez-Gil, Agustín Valenzuela-Fernández, Rafaela González-Montelongo, Laura Ciuffreda, Carlos Flores

## Abstract

**Objectives:** Limited testing capacity has characterized the ongoing COVID-19 pandemic in Spain, hampering a timely control of outbreaks and the possibilities to reduce the escalation of community transmissions. Here we investigated the potential of using pooling of samples followed by one-step retrotranscription and quantitative PCR (RT-qPCR) to increase SARS-CoV-2 testing capacity.

**Methods:** We first evaluated different sample pooling (1:5, 1:10 and 1:15) prior to RNA extractions followed by standard RT-qPCR for SARS-CoV-2/COVID-19 diagnosis. The pool size achieving reproducible results in independent tests was then used for assessing nasopharyngeal samples in a tertiary hospital during August 2020.

**Results:** We found that pool size of five samples achieved the highest sensitivity compared to pool sizes of 10 and 15, showing a mean (± SD) Ct shift of 3.5 ± 2.2 between the pooled test and positive samples in the pool. We then used a pool size of five to test a total of 895 pools (4,475 prospective samples) using two different RT-qPCR kits available at that time. The Real Accurate Quadruplex corona-plus PCR Kit (PathoFinder) reported the lowest mean Ct (± SD) shift (2.2 ± 2.4) among the pool and the individual samples. The strategy allows detecting individual samples in the positive pools with Cts in the range of 16.7-39.4.

**Conclusions:** We found that pools of five samples combined with RT-qPCR solutions helped to increase SARS-CoV-2 testing capacity with minimal loss of sensitivity compared to that resulting from testing the samples independently.

## Introduction

The SARS-CoV-2 pandemic causing COVID-19 continue imposing a heavy burden on healthcare systems worldwide because of a shortage of consumables and the demand for scaling up efficient screening approaches. To limit the escalation of cases and amplification of infections, increasing the capacities and developing alternatives to the one-step reverse transcription and real-time quantitative PCR (RT-qPCR) for regular testing of SARS-CoV-2 is key (Mina *et al*. 2020). We have assessed a direct heating method of nasopharyngeal (swab) samples to bypass the RNA extraction step for increasing the testing capacity (Alcoba-Florez *et al*. 2020a,b).

In areas with low COVID-19 prevalence, such as the Canary Islands until June 2020 (Pollán *et al*. 2020), testing of samples in pools is another approach that can efficiently increase SARS-CoV-2 testing capacity. Simulation studies support the value of testing on sample pools and the speed of reporting, while the impact of the sensitivity of tests is comparable smaller (Larremore *et al*. 2020). Supporting this, a pooled test of SARS-CoV-2 of four samples has received Emergency Use Authorization from the USA Food and Drug Administration (FDA COVID-19 Update 2020).

Here we aimed to evaluate the RT-qPCR testing on pooled swab samples with the goal to demonstrate its feasibility as an option to increase SARS-CoV-2 testing capacity in the region.

## Materials and Methods

The study was conducted in two stages at the University Hospital Nuestra Señora de Candelaria (Santa Cruz de Tenerife, Spain) during August 2020. Nasopharyngeal swab samples were collected in 2 mL of viral transmission medium (VTM) (Biomérieux).

RNA extractions were conducted from 200 μL of pooled VTMs using the MagNA Pure Compact Nucleic Acid Isolation Kit I (Roche) or the STARMag Viral DNA/RNA 200C kit (Seegene) as described elsewhere (Alcoba-Florez *et al*. 2020a,b). Two RT-qPCR solutions were used in the study period: The Real Accurate Quadruplex corona-plus PCR Kit (PathoFinder), and the TaqPath COVID-19 CE-IVD RT-PCR Kit (Thermo Fisher Scientific). We focused on the results for the N target gene for both kits (Alcoba-Florez *et al*. 2020b). The RT-qPCR was performed in 10 μL final volume reactions (5 μL of sample) using a CFX96 Touch Real-Time PCR Detection System (Bio-Rad) following the thermal cycling specifications of each solution. Samples were considered negative when the SARS-CoV-2 target had Ct >40. Positive and negative controls were included in all experiments as described elsewhere (Alcoba-Florez *et al*. 2020a,b).

In a pilot stage, we used the Real Accurate Quadruplex corona-plus PCR Kit for testing 15 pools made by combining 5, 10, or 15 retrospective samples, each containing equal volumes of a SARS-CoV-2/COVID-19 positive sample and the respective amounts of negative samples to complete the pool size. In a validation stage, we used the two RT-qPCR kits available on swab samples from prospective subjects using the pooling size achieving optimal results. Then, individual samples from each positive pool were subjected to RNA extraction followed by RT-qPCR using the above-mentioned methods to validate the results.

Differences between the pooled Ct and the positive sample Ct in the pool (Ct shift) for the two different RT-qPCR kits were assessed by Mann-Whitney U-test using the R v4.0.3 software. When more than a positive sample was present in the pool, the mean Ct of the positive samples was used in the calculation. Sensitivity and the 95% Confidence Interval (CI) was assessed with MedCalc (MedCalc Software Ltd.).

## Results

In a pilot study, detection of SARS-CoV-2 was achieved with pool size of five (one positive) in 13 out of 15 independent pools (sensitivity = 86.7% [95% CI= 59.5-98.3]), where detected positives had a maximum Ct value of 36.2 (**Figure 1**). A mean (± SD) Ct shift of 3.5 (± 2.2) was obtained between the pool of five samples and the individual positive samples. False negatives increased for pool sizes of 10 and 15 samples, both showing larger mean Ct shifts (5.3 ± 2.4 and 7.2 ± 4.4, respectively).

**Figure 1.**
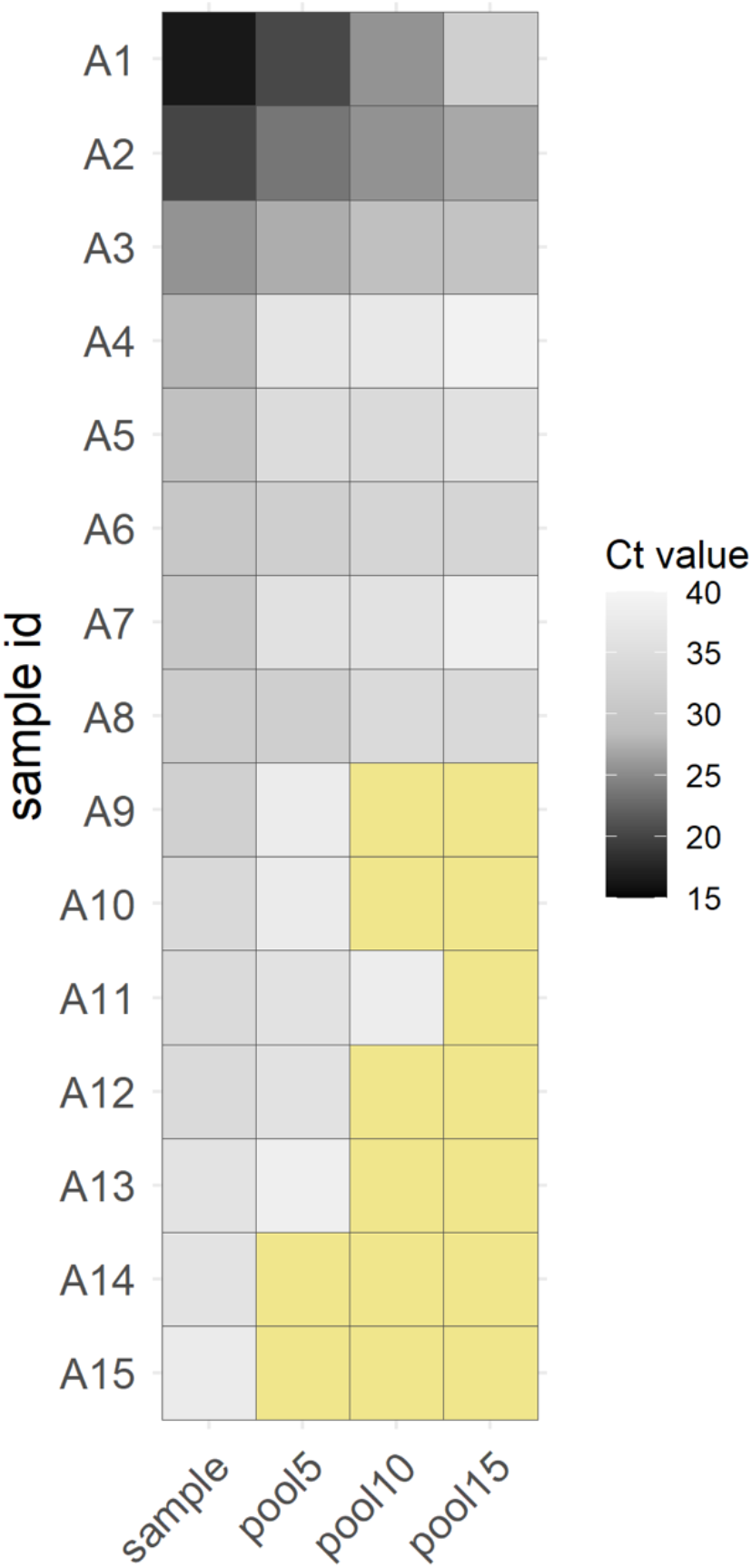
Heatmap representation of RT-qPCR test positiveness on a Ct scale (the darker, the lower the Ct) on the pilot study with 1:5 (pool 5), 1:10 (pool 10), and 1:15 (pool 15) pooling of retrospective swab samples. Negative results (Ct>40) are highlighted in pale yellow. The Real Accurate Quadruplex corona-plus PCR Kit (PathoFinder) was used in the experiment.

A validation study included samples from 4,475 prospective subjects between August 18^th^ 2020 and August 31^st^ 2020. Samples were tested in 546 pools (447 negatives) for the Real Accurate Quadruplex corona-plus PCR Kit and 349 (286 negatives) for the TaqPath COVID-19 CE-IVD RT-PCR Kit. A total of 162 pools were tested positive among the two kits, 118 containing just one positive sample and 44 containing more than one positive sample (**Figure 2**). Compared to the independent samples present in the positive pools, the mean Ct shift upon pooling was 2.2 ± 2.4 for the Real Accurate Quadruplex corona-plus PCR Kit and 3.1 ± 2.9 for the TaqPath COVID-19 CE-IVD RT-PCR Kit (*p*=0.006, Mann-Whitney U-test). Both RT-qPCR kits allowed to sensitively detect positive samples with a maximum Ct value of 39.4.

**Figure 2.**
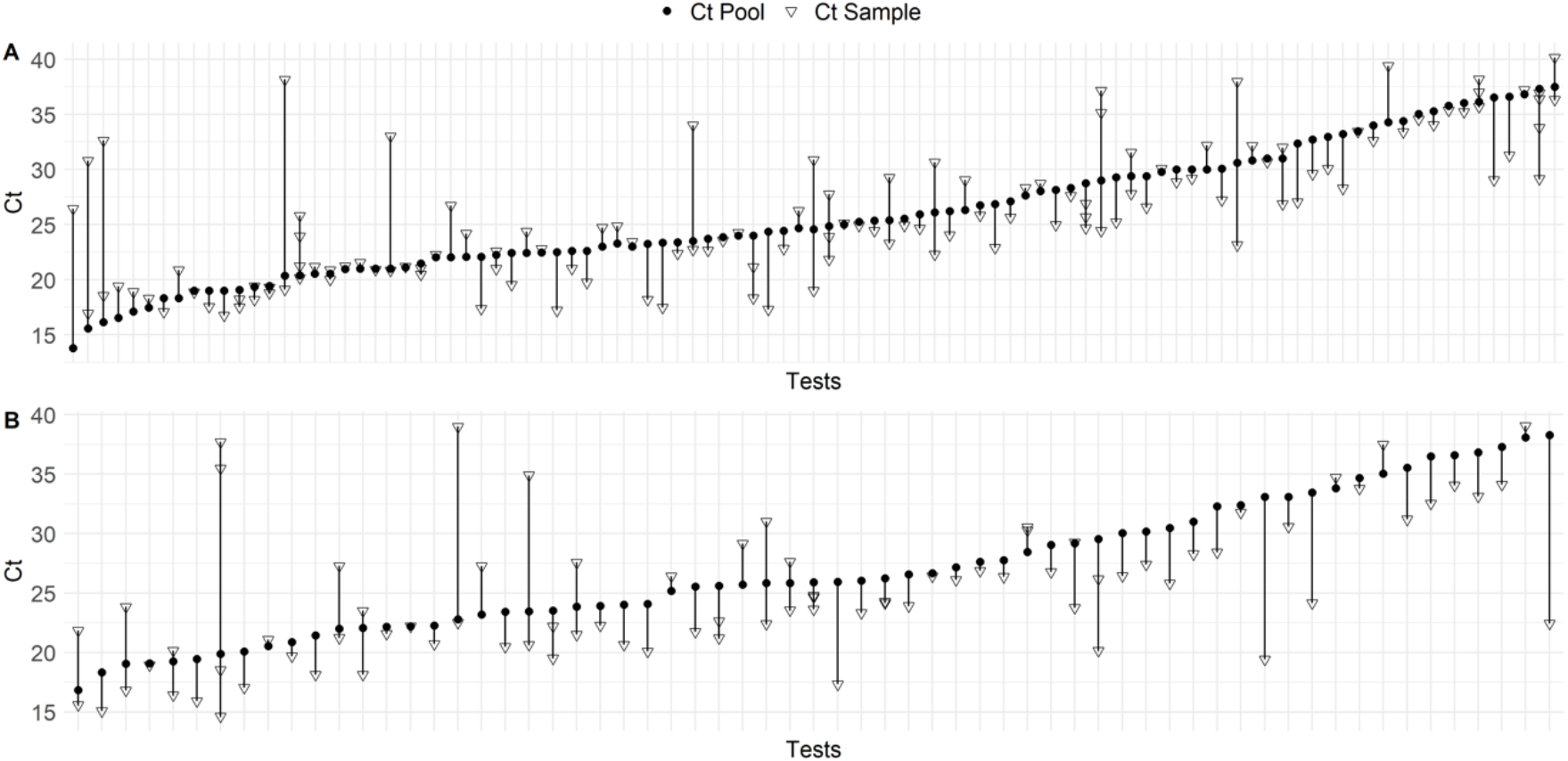
Scatter plot showing Ct values and Ct shifts for all prospective COVID-19 positive cases detected by RT-qPCR on 1:5 pooling tests (full circle) and on individual samples (empty triangle) using (**A**) the Real Accurate Quadruplex corona-plus PCR Kit and (**B**) the TaqPath COVID-19 CE-IVD RT-PCR Kit.

## Discussion

In this study, we describe the feasibility of RT-qPCR in pools of five swab samples with a minimal loss of sensitivity compared to the assessment of samples independently. Consistent with these findings, others have recently demonstrated that pooling of up to 10 samples result in a slight shift in Ct (around 3), and therefore a drop of sensitivity (Das *et al*. 2020).

A major limitation of the study is that sensitivity was only evaluated in the pilot stage. However, despite the small Ct shift between the positive pools and the individual positive samples in the prospective study, a high impact of false negatives is not expected (Cherif *et al*. 2020). Our data suggest that our sample pooling strategy is able to detect COVID-19 positive cases with Ct values ≤39.4. Pooling with compressive sampling designs, which involves repetitive tests (Shental *et al*. 2020), and the use of alternative testing approximations (Peto *et al*. 2020) may help to further lessen the impact of the dilution factor on pooling and continue increasing health capacity building.

## Data Availability

The data that support the findings of this study are available on request from the corresponding author.

## Authors’ contributions

JAF and CF designed the study. JAF, HGC, DGM, and ODG participated in data acquisition. JAF, LC, and CF performed the analyses and data interpretation. LC, AVF, RGM and CF wrote the draft of the manuscript. All authors contributed in the critical revision and final approval of the manuscript.

## Acknowledgments

We deeply acknowledge the University Hospital Nuestra Señora de Candelaria board of directors and the executive team for their strong support and assistance in accessing diverse resources used in the study.

## Ethical Approval

The University Hospital Nuestra Señora de Candelaria (Santa Cruz de Tenerife, Spain) review board approved the study (ethics approval number: CHUNSC_2020_24).

## References

Alcoba-Florez J, González-Montelongo R, Íñigo-Campos A, García-Martínez de Artola D, Gil-Campesino H, The Microbiology Technical Support Team, Ciuffreda L, Valenzuela-Fernández A, Flores C. Fast SARS-CoV-2 detection by RT-qPCR in preheated nasopharyngeal swab samples. Int J Infect Dis. 2020a May 31;97:66–68.

Alcoba-Florez J, Gil-Campesino H, García-Martínez de Artola D, González-Montelongo R, Valenzuela-Fernández A, Ciuffreda L, Flores C. Sensitivity of different RT-qPCR solutions for SARS-CoV-2 detection. Int J Infect Dis. 2020b Jul 31;99:190–192.

Cherif A, Grobe N, Wang X, Kotanko P. Simulation of pool testing to identify patients with coronavirus disease 2019 under conditions of limited test availability. JAMA Netw Open. 2020 Jun 23; 3:e2013075.

FDA COVID-19 Update: FDA Issues First Emergency Authorization for Sample Pooling in Diagnostic Testing Jul 18, 2020. https://www.fda.gov/news-events/press-announcements/coronavirus-covid-19-update-fda-issues-first-emergency-authorization-sample-pooling-diagnostic

Das S, Lau AF, Youn JH, Khil PP, Zelazny AM, Frank KM. Pooled testing for surveillance of SARS-CoV-2 in asymptomatic individuals. J Clin Virol. 2020 Sep 3; 132:104619.

Larremore DB, Wilder B, Lester E, Shehata S, Burke JM, Hay JA, Tambe M, Mina MJ, Parker R. Test sensitivity is secondary to frequency and turnaround time for COVID-19 surveillance. MedRxiv 2020 Sept 8. doi: 10.1101/2020.06.22.20136309.

Mina MJ, Parker R, Larremore DB. Rethinking Covid-19 Test Sensitivity - A Strategy for Containment. N Engl J Med. 2020 Sep 30. doi: 10.1056/NEJMp2025631.

Peto L, Rodger G, Carter DP, Osman KL, Yavuz M, Johnson K, Raza M, Parker MD, Wyles MD, Andersson M, Justice A, Vaughan A, Hoosdally S, Stoesser N, Matthews PC, Eyre DW, Peto TEA, Carroll MW, de Silva TI, Crook DW, Evans CM, Pullan ST. Diagnosis of SARS-CoV-2 infection with LamPORE, a high-throughput platform combining loop-mediated isothermal amplification and nanopore sequencing. MedRxiv. 2020 Sep 25. doi: 10.1101/2020.09.18.20195370.

Pollán M, Pérez-Gómez B, Pastor-Barriuso R, Oteo J, Hernán MA, Pérez-Olmeda M, Sanmartín JL, Fernández-García A, Cruz I, Fernández de Larrea N, Molina M, Rodríguez-Cabrera F, Martín M, Merino-Amador P, León Paniagua J, Muñoz-Montalvo JF, Blanco F, Yotti R; ENE-COVID Study Group. Prevalence of SARS-CoV-2 in Spain (ENE-COVID): a nationwide, population-based seroepidemiological study. Lancet. 2020 Aug 22; 396:535–544.

Shental N, Levy S, Skorniakov S, Wuvshet V, Shemer-Avni Y, Porgador A, Hertz T. Efficient high throughput SARS-CoV-2 testing to detect asymptomatic carriers. MedRxiv 2020 Apr 20. doi:10.1101/2020.04.14.20064618.

